# Pilot screening multivariable prediction model for late diagnosed cystic fibrosis

**DOI:** 10.64898/2026.02.09.26343980

**Authors:** Richard A Belkin, Yuri Matusov, Barry Belkin

**Affiliations:** Department of Pulmonary and Critical Care Medicine, Santa Barbara Cottage Hospital, Santa Barbara, CA, United States of America; Division of Pulmonary and Critical Care Medicine, Department of Medicine, Cedars-Sinai Medical Center, Los Angeles, CA, United States of America; Independent mathematician/statistician, Santa Barbara, CA, United States of America

**Keywords:** Screening, Algorithm, Prediction, Adult Cystic Fibrosis

## Abstract

**Background:** Diagnosing cystic fibrosis (CF) in adults is challenging. Patients often present with atypical and varied symptoms. The aim of this study is to develop a pilot prediction model to aid clinicians in identifying patients who should undergo appropriate CF diagnostic testing.

**Methods:** This is a retrospective, single-center, observational cohort study of adult (≥18 yrs) patients who underwent sweat testing for evaluation of clinically suspected CF. Dataset 1 includes 171 patients with CF-supportive conditions from 2005-2021 and was used both to develop the screening model and to subject it to internal validation. Dataset 2 includes 87 patients with CF-supportive conditions from 2021-2023 and was used to subject the prediction model to external validation.

Based on CF Foundation guidelines, dataset patients were each assigned one of three CF classifications: (1) CF likely (Type *A*), (2) CF not resolved (Type *B*), or (3) CF unlikely (Type *C*). Clinical predictor variables indicate the presence (value=1) or absence (value=0) of 7 CF-related clinically observable patient conditions. The screening model decision rule recommends a patient for CF testing if either of two conditions holds: (1) A numerical patient score *S*_*A*_ calculated using multivariate logistic regression (LR) equals or exceeds a threshold value *C*_0_ determined using ROC curve analysis, or (2) the patient diagnosis is positive for bronchiectasis/tree-in-bud and negative or unknown for each of 6 other clinical conditions.

**Results:** For each dataset, the observed sensitivity value for recommending CF testing was 1.00 for patients that CF testing classified as Type *A* or Type *B*. Further, for the two datasets in combination, 2/128=1.56% of the Type *C* patients were not recommended for CF testing.

**Conclusions:** When applied separately to each dataset, the screening model matched the predictive accuracy of clinical judgment both for Type *A* and Type *B* patients. A similar predictive accuracy comparison for Type *C* patients is not possible because the CF types are unknown for patients not recommended for CF testing.

## 1. Introduction

Adult diagnosed CF and CFTR-RD patients tend to have CFTR variants with residual CFTR function, may have intermediate (or low) sweat chloride values, and often present with a more atypical clinical phenotype than that of the childhood CF presentation.(1,2) CF is diagnosed when an individual has both a clinical presentation of the disease and evidence of CFTR dysfunction based on CF Foundation (CFF) guidelines, whereas CFTR-RD has been defined as a clinical condition with evidence of CFTR protein dysfunction that does not fulfill the diagnostic criteria for CF.(3,4) Diagnosing CF is particularly challenging when the adult patient presents with unexplained chronic sinopulmonary disease, recurrent pancreatitis, or infertility. The adult patient symptoms and findings may be so insidious that a diagnosis of CF is not considered even by an experienced clinician.(3) The most significant benefits of an accurate diagnosis of CF are access to highly effective CFTR modulator therapy, now available for approximately 90% of patients, and multidisciplinary care at an accredited CF center. In addition, having an established diagnosis allows for access by the patient and family members to genetic counseling, when reproduction is being considered, and relief of diagnostic uncertainty.(4,5) It is also important to minimize CF testing where CF is unlikely in order to decrease cost and patient anxiety that may develop when CF testing is ordered.

To address the multiple challenges of adult CF diagnosis, an active screening program for sweat chloride testing was initiated at our CF center in 2005 through outreach and education of our medical community.

The present study has two primary objectives: The first is to design a pilot multivariable prediction model whose function is to recommend whether a patient should undergo diagnostic laboratory testing for CF (sweat chloride testing, and when indicated, subsequent CFTR mutation testing). The second objective was to test the accuracy of the predictive model when applied to patient data.

## 2. Materials and Methods

The study was approved by the Santa Barbara Cottage Hospital Institutional Review Board (IRB #16-94) on 2/15/2017. The IRB approved waivers of consent and HIPAA authorization.

### 2.1 Study type, sources of data, participants, and outcome

This is a single-center, retrospective, observational study of adult patients (≥18 years old) who were: (i) screened for CF by affiliated physicians, predominantly pulmonologists and gastroenterologists, using a sweat chloride test at Santa Barbara Cottage Hospital (SBCH), or (ii) referred to and evaluated at the Cottage Cystic Fibrosis and Bronchiectasis Center, with a diagnosis of CF under consideration and with at least one completed sweat chloride test. The Cottage Cystic Fibrosis and Bronchiectasis Center is a CFF-accredited, multidisciplinary CF program affiliated with a community teaching hospital and functions as a tertiary referral center both for patients with newly diagnosed adult CF and patients for whom the diagnosis is being considered. The patients included in the study were screened for CF via sweat chloride testing between January 2005 and December 2023. Age listed for each patient was based on the date the first sweat test was performed.

The type of prediction model we used was nonrandom split-sample development and validation based on TRIPOD guidelines.(6) The initial study population (Dataset 1) comprised of 171 patients tested for CF between January 2005 and April 2021. This dataset was used both in developing and training the prediction model, and in the internal validation of the prediction model. A second study population (Dataset 2) comprised of 87 patients tested for CF between May 2021 and December 2023 and was used in the validation of the prediction model. Patients were excluded from the study if insufficient clinical data relevant to patient screening for CF testing were available. The screening function of the prediction model is to determine whether the available clinical evidence supports a recommendation that the patient should be tested for CF.

Data for this study were accessed between February 15, 2017 and January 2, 2025. Yuri Matusov, MD, had access to identifying information for data collection from February 15, 2017 to June 30, 2018. Richard Belkin, MD, had access to identifying information throughout the entire course of data collection (February 15, 2017 to January 2, 2025). Richard Belkin, MD, is the only author who continues to have access to identifying information after completion of data collection.

### 2.2 Methods Overview

The basic analysis framework for the screening model is described below. The Supplement provides detailed explanatory material together with supporting mathematical and statistical detail.

Based on CF testing as outlined in Figure 1 and after thorough clinical chart review, patients were classified by the CF center medical director (RB) as follows: Type *A* (CF likely), Type *B* (CF not resolved (CFNR), including CFTR-RD), or Type *C* (CF unlikely). This CF classification scheme is based on the CFF diagnosis guidelines (7–9) specified in Figure 1.

**Fig 1:**
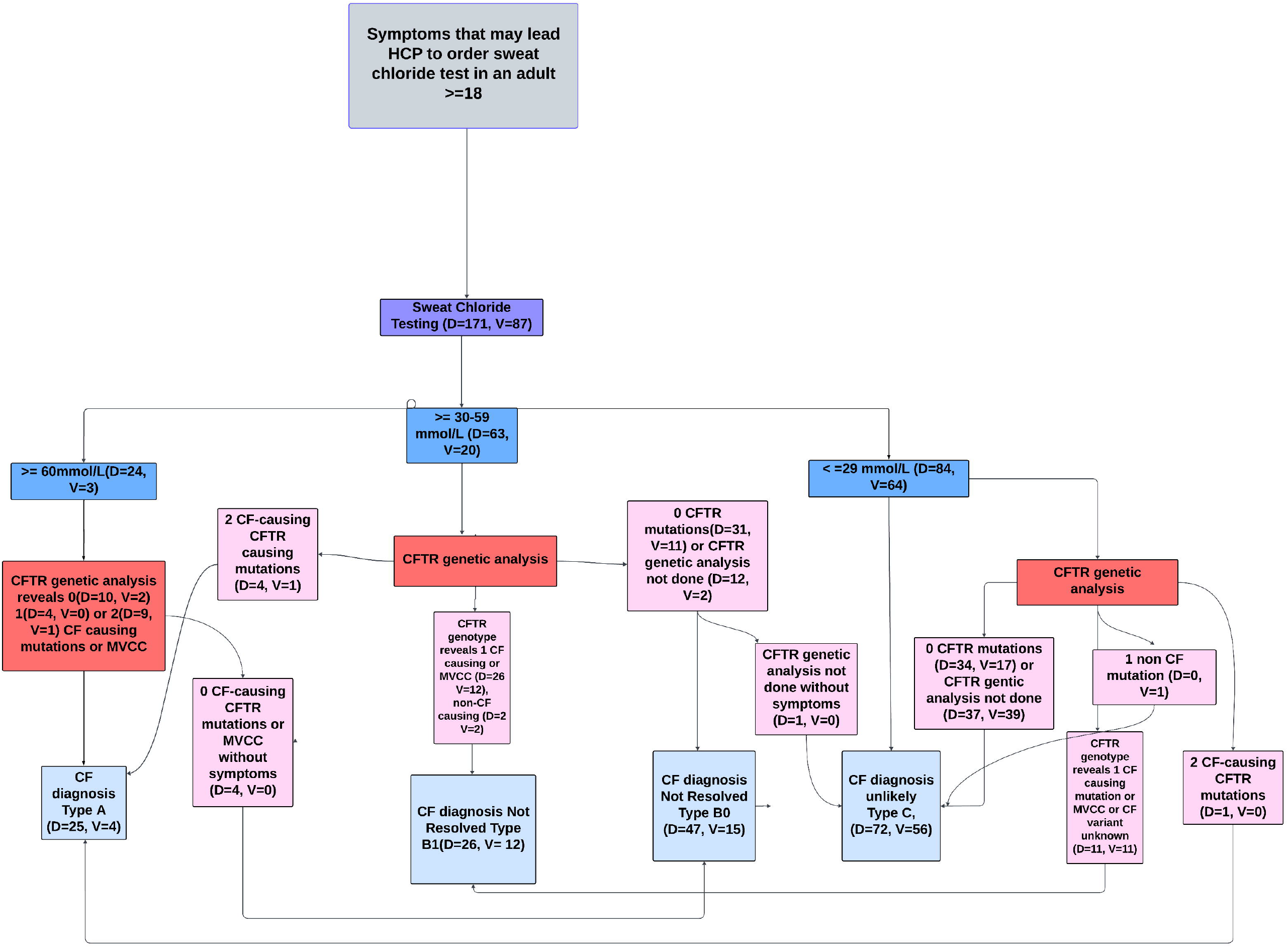
Flowchart for diagnosis of CF and CFNR (includes patients with CFTR-RD). D=Development cohort (Dataset 1) and V=External Validation cohort (Dataset 2). Diagnosis of CF based on CF Foundation diagnosis guidelines.(7,8) Of note, our CF center does not have routine access to nasal potential difference (NPD) or intestinal current measurement testing to assist in CF diagnoses.

Type *B* was further separated into two subgroups: (1) patients (designated Type *B*_1_) with one CFTR gene variant or non-CF causing variant, including CF and VVCC and (2) patients (designated Type *B*_0_) who were either (a) reported with no CFTR gene variant or (b) not tested for CFTR gene variants (based on the sweat test results and the judgment of the clinician). See Table S3-D and S3-V for breakdown of Type *B* patients. The importance of testing and identifying Type *B*_1_ patients for CF lies in the potential candidacy of a Type *B*_1_ patient for CFTR modulator or other genetic therapies for CF. Although the testing and identifying of Type *B*_0_ patients maybe of lesser importance than the testing and identifying of Type *B*_1_ patients, the intended performance goal for the screening model is to recommend for CF testing all Type *A*, Type *B*_1_, and Type *B*_0_ patients. *A, B*_1_, and *B*_0_ patients recommended for CF testing by the prediction model are counted as true positives. Type *C* patients recommended for CF testing by the screening model are counted as false positives. See the attached diagnosis flowchart, Figure 1.

### 2.3 Clinical predictors

The study data included 23 potential variables (Supplement Table S4-D and S4-V) referred to as *clinical predictors*, each of which records the presence (value=1), absence (value=0), or the unknown diagnosis of a specific clinical condition known to be observed in Type *A*. Candidate clinical predictors were chosen based on prior knowledge of their presence in adult diagnosed CF patients and on their potential to be predictive of CF. Patient data for the particular clinical predictors selected for possible use in screening for CF testing was extracted from retrospective electronic chart review using EPIC (Epic Systems, Verona, WI) and Elation (Elation Health, San Francisco, CA). These data were managed using RED Cap electronic data capture tools hosted at SBCH (10,11) and MS Excel (Version 16.0.14931.20132, Redmond, WA).

A given clinical condition was determined to be present in a patient if it was present on chart review at any timepoint either before or after sweat testing was done. BMI and FEV1 were collected at the timepoint closest to when sweat testing was performed. Pancreatic insufficiency was determined to be present if listed in the patient’s clinical chart or if the patient had a low pancreatic fecal elastase level (<500 µg/g). The presence of bronchiectasis and tree-in-bud opacities was identified either in a chest CT report or in the problem list/assessment, if CT reports were not available. Other variables such as acute or chronic pancreatitis were determined to be present based on detailed review of the medical chart. See dataset repository for the list of clinical predictors considered in the prediction model.

Importance testing based on statistical metrics was used to reduce the set of 23 candidate clinical conditions under consideration to the subset of 15 clinical conditions used in the predictive model. One of the two metrics, the odds ratio (OR) measures the degree to which knowledge that a patient is diagnosed as positive for a specified CF-related clinical condition increases the probability that the patient CF classification is Type *A*. For a specified CF-related clinical condition, the OR is given by the following ratio:

Fraction of Dataset 1 patients positive for the CF-related clinical condition who are Type *A* Fraction of Dataset 1 patients who are Type *A*

The second metric measures the likelihood (prevalence) of a specified CF-related clinical condition in patients classified as Type *A*.

In some cases, composites of multiple candidate clinical conditions were formed, such that a patient is positive for the composite condition if positive for one or more of the associated component conditions. This reduced the final set of clinical conditions used in the screening model from 15 clinical conditions to the 7 clinical conditions or composite clinical conditions listed in Table 1.

### 2.4 Statistical Analysis

The screening model decision rule recommends a patient for CF testing if either one or both of the following requirements are satisfied: (Criterion 1) a numerical patient score *S*_*A*_ with a value between 0 and 1 equals or exceeds a threshold value *C*_0_ calculated using logistic regression (LR) and ROC curve analysis; (Criterion 2) the patient diagnosis is positive for bronchiectasis/tree-in-bud and either negative or unknown for the remaining 6 CF-related clinical conditions.

The value of *S*_*A*_ for a given patient quantifies the weight of clinical evidence supporting a Type *A* classification and is determined using LR analysis, as explained in Section S.1 of the Supplement.

As reported in the Results Section below, for all Dataset 1 and Dataset 2 patients classified as Type *A* patients, Criterion 1 is satisfied. As a result, all Type *A* patients are recommended for CF testing by the screening model. However, it is not the case that Criterion 1 is satisfied for all of the Type *B* patients. The LR analysis results indicate that all of the Type *B* patients not recommended for CF testing on the basis of Criterion (1) also have bronchiectasis/tree-in-bud present and either a negative or unknown diagnosis for each of the 6 remaining CF-related clinical conditions. Thus, criterion (2) is met, and all Type *B* patients are recommended for CF testing by the screening model. It is also the case that criterion (2) is satisfied by some Type *C* patients.

### 2.5 Use of multiple LR models

Of the 7 CF-related clinical conditions listed in Table 1 selected for use in the LR analysis, the average number for which a Dataset 1 patient is positive is 2.01. Of the 171 Dataset 1 patients, only 16 patients are positive for more than 3 of the selected clinical conditions. The set of 7 selected clinical conditions has 35 subsets of size 3. The approach taken then is that LR analysis with 3 clinical predictors is separately applied to each of these 35 subsets of size 3. For each patient, each of the 35 LR models produces an estimate for the probability that the patient CF classification is Type *A*. The value of the score *S*_*A*_ for the patient is then taken to be the largest of these 35 probability estimates.

**Table 1.**
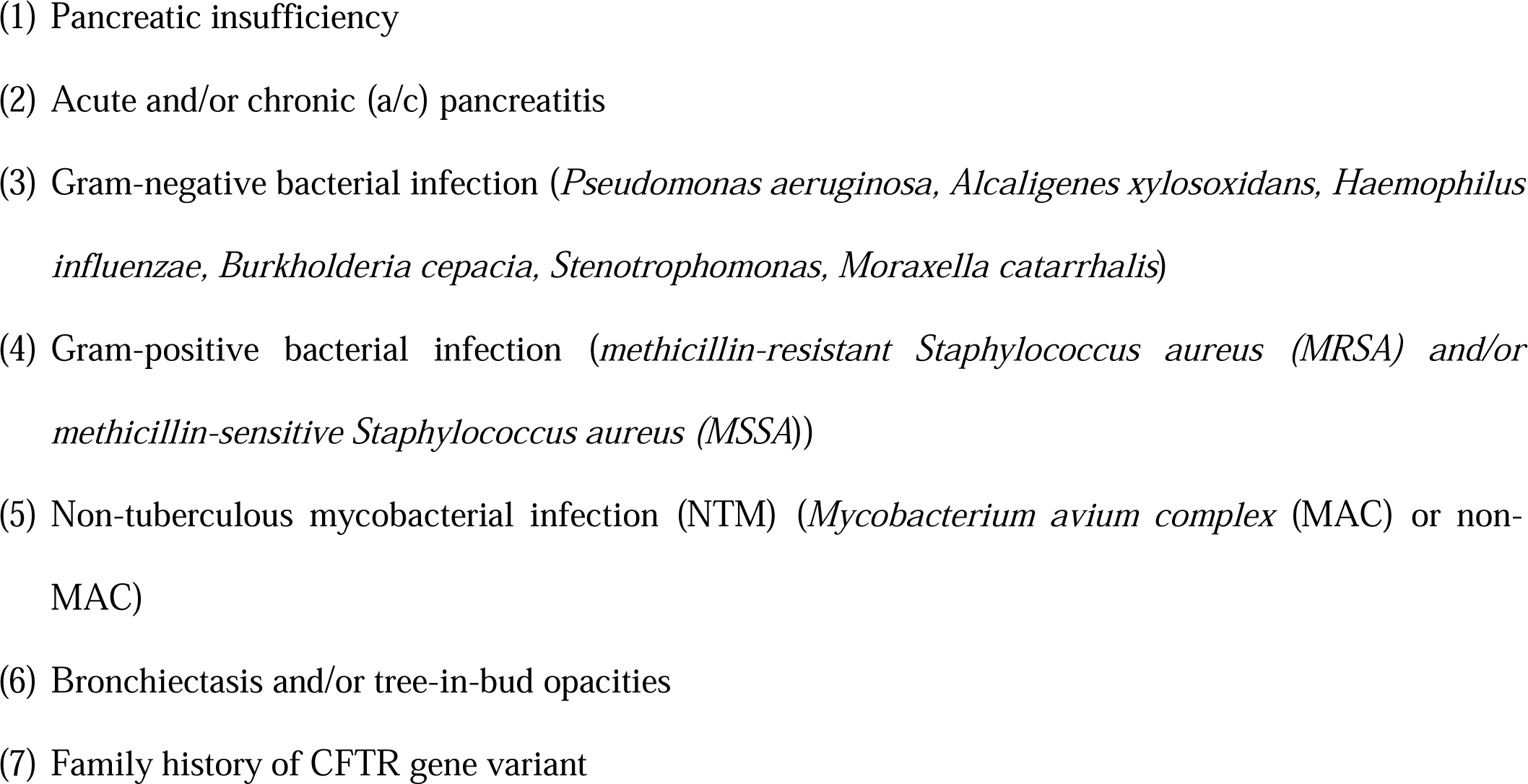
List of 7 CF-Related Clinical Conditions or Composite Clinical Conditions Selected for Use in LR Analysis.

The decision based on LR analysis whether to recommend a patient for CF testing is thus based on the particular set of 3 clinical predictors that provides the strongest clinical evidence that the patient CF classification is Type *A*.

### 2.6 LR model calibration

The values of the parameters for each of the 35 different 3-variate LR models are determined from Dataset 1 as described in Section S.2 of the Supplement. An important feature of the LR model calibration is that the data for all 171 Dataset 1 patients are used in calibrating each of the 35 separate 3-variate LR models. This multiple model approach to the LR analysis addresses the problem of model overfitting that would be present if for each patient, *S*_*A*_ were based on a single 7-variate LR model.

### 2.7 LR decision rule for recommendation of CF testing

The value of *S*_*A*_ is compared with a threshold *C*_0_ whose value is determined using ROC curve analysis. The decision rule used by the screening algorithm is: if *S*_*A*_ ≥ *C*_0_, then CF testing is recommended; if *S*_*A*_ < *C*_0_, then CF testing is not recommended.

### 2.8 Choice of value for *C*_0_

An important subject of study investigation is whether the value of *C*_0_ can be chosen so that if the screening algorithm is applied to Datasets 1 and 2, in both cases, two conditions are met: (1) the calculated sensitivity value is 1 for recommending Type *A* patients for CF testing and (2) the calculated sensitivity value is 1 for recommending Type *B* patients for CF testing.

### 2.9 Missing data

Patients for whom most of the clinical information was missing were excluded from the study. This applied to a very small number of patients. If a specific clinical condition was missing for an individual patient, the clinical condition was labelled as unknown in the LR models used for development and validation. If a clinical condition or composite clinical condition was missing upon chart review, it was either labelled as not present if the CF program director (RB) felt it was unlikely that the associated clinical condition was present or left as unknown if considered to be truly unknown (e.g., if the patient had (1) no respiratory symptoms and a normal chest x-ray or CT scan of the chest, and (2) no cultures were performed, it was assumed that no gram postive, gram negative or mycobacterial infections were present). The number of predictors used in the LR analysis is adjusted to be equal to the number of CF-related clinical conditions for which there is data. For example, in cases where the value of one of the predictor variables in a 3-variate LR model wwas truly unknown for a given patient, a bivariate LR model was substituted for 3-variate LR.

### 2.10 Predictive Accuracy of Screening Model

The results of separately applying the screening model to each dataset are in the form of three metrics: sensitivity values for Type *A* and Type *B*, and a specificity value for Type *C*. Consideration was given to the use of the bootstrap method of randomized resampling as a means of quantifying the estimation errors associated with these metrics. However, as indicated in the Results section, for both Dataset 1 and Dataset 2, the sensitivity values for Type *A* and Type *B* are each 1.00. So, there is no statistical variation for the bootstrapping method to quantify. The testing of the predictive accuracy of the screening model is further described in Section S.3 of the Supplement.

### 2.11 Illustrative example of application of screening model

The application of the screening model to a patient is described in detail in Section S.4 of the Supplement. An illustrative example of the application of the screening model on a patient not included in our study datasets is provided in the Supplement Section S.5.

## 3. Results

### 3.1 Participants

The baseline demographics and clinical characteristics can be found in Tables S1-5D and S1-5V. In the 171-patient development cohort, 25 patients (15%) were classified as having CF, 74 (43%) were classified as CFNR, and 72 (42%) were classified as not having CF or CF-related disease. These patients had a median age of 53, 61, and 58 years, respectively, and all three groups had female predominance (70% of the development cohort). Of the CF group, 20 (80%) had sweat chloride > 60 mmol/L, whereas 58 (78%) of the CFNR group had sweat chloride between 30-59 mmol/L (refer to Table S1-D). In the 87-patient validation cohort, 4 (5%) were classified as having CF, 27 (31%) were classified as CFNR, and 56 (64%) were classified as CF unlikely. These patients had a median age of 59 (D) and 62 (V) years and had a female predominance with a similar distribution of sweat chloride levels (refer to Table S1-V).

### 3.2 Selection of CF-related clinical conditions and model development with Dataset 1

As previously described, the screening algorithm is based on multiple 3-variate LR models. As the number of CF-related clinical conditions considered in the LR analysis increases, so does the number of different 3-variate LR models. With 7 CF-related clinical conditions, the number of different 3-variate LR models is 35 (the number of subsets with 3 elements of a set with 7 elements).

Seven CF-related clinical conditions were ultimately chosen for analysis and modeling (Table 2). Patients were more likely to have Type *A* if they had pancreatic insufficiency (OR 4.56), acute/chronic pancreatitis (OR 2.49), a family history of CFTR gene variants (OR=2.41), and respiratory tract gram positive bacterial infection (OR=1.55).

**Table 2.** Observed Odds Ratios and Prevalence Values for the 7 Selected CF-Related Clinical Conditions.

| The 7 Selected CF-Related Clinical Conditions | Odds Ratio (OR) | Prevalence |
| --- | --- | --- |
| Pancreatic Insufficiency | 4.56 | 0.07 |
| Acute/Recurrent/Chronic Pancreatitis | 2.49 | 0.06 |
| Family History of CFTR Gene Variant | 2.41 | 0.10 |
| Respiratory Tract Gram-Negative Bacterial Infection | 1.02 | 0.51 |
| Respiratory Tract Gram-Positive Bacterial Infection | 1.55 | 0.31 |
| NTM (MAC or non-MAC) | 0.70 | 0.29 |
| Bronchiectasis and/or Tree-in-Bud Opacities | 0.70 | 0.67 |

The results that follow were obtained by applying the multivariate LR methods described with the rule that the patients referred for CF testing are those for whom the inequality *S*_*A*_ ≥ *C*_0_ is satisfied.

Table 3 lists, in decreasing order of AUC value, the combinations of 3 CF-related clinical conditions with AUC values > 0.7. Each of the 7 selected CF-related clinical conditions appears in Table 3 at least once. Application of the screening algorithm to the development cohort showed the highest AUC value for the combination of pancreatic insufficiency, acute/chronic pancreatitis, and bronchiectasis or tree-in-bud opacities (AUC=0.786). This was closely followed by the combination of pancreatic insufficiency, bronchiectasis or tree-in-bud opacities, and respiratory tract gram-negative bacterial infection (AUC=0.770). Application of the screening algorithm to the 171-patient development cohort resulted in the ROC curve shown in Figure 2, with an AUC value of 0.829.

**Table 3.**
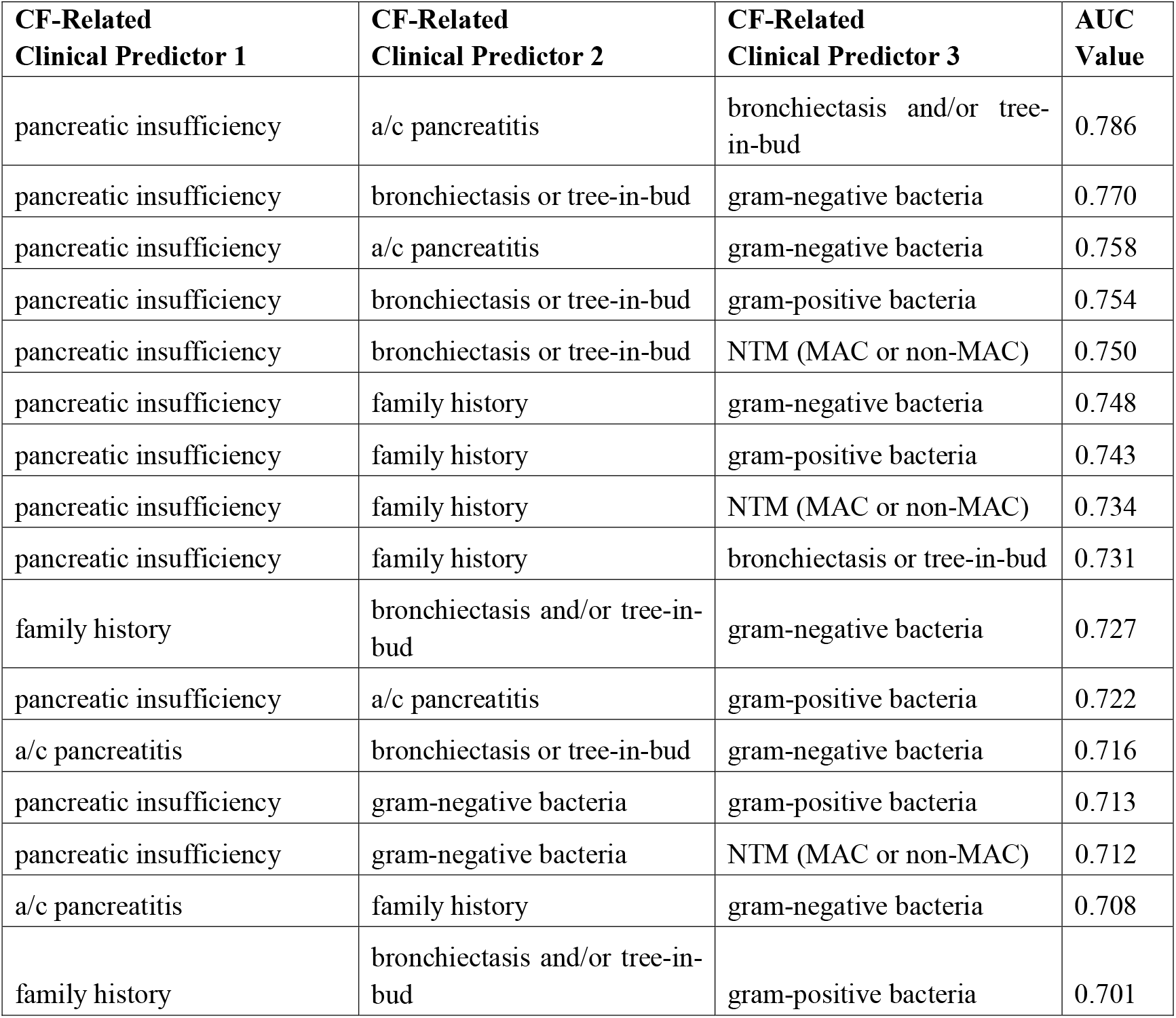
List of 3-variate LR models with AUC Values in Excess of 0.700.

| CF-Related Clinical Predictor 1 | CF-Related Clinical Predictor 2 | CF-Related Clinical Predictor 3 | AUC Value |
| --- | --- | --- | --- |
| pancreatic insufficiency | a/c pancreatitis | bronchiectasis and/or tree-in-bud | 0.786 |
| pancreatic insufficiency | bronchiectasis or tree-in-bud | gram-negative bacteria | 0.770 |
| pancreatic insufficiency | a/c pancreatitis | gram-negative bacteria | 0.758 |
| pancreatic insufficiency | bronchiectasis or tree-in-bud | gram-positive bacteria | 0.754 |
| pancreatic insufficiency | bronchiectasis or tree-in-bud | NTM (MAC or non-MAC) | 0.750 |
| pancreatic insufficiency | family history | gram-negative bacteria | 0.748 |
| pancreatic insufficiency | family history | gram-positive bacteria | 0.743 |
| pancreatic insufficiency | family history | NTM (MAC or non-MAC) | 0.734 |
| pancreatic insufficiency | family history | bronchiectasis or tree-in-bud | 0.731 |
| family history | bronchiectasis and/or tree-in-bud | gram-negative bacteria | 0.727 |
| pancreatic insufficiency | a/c pancreatitis | gram-positive bacteria | 0.722 |
| a/c pancreatitis | bronchiectasis or tree-in-bud | gram-negative bacteria | 0.716 |
| pancreatic insufficiency | gram-negative bacteria | gram-positive bacteria | 0.713 |
| pancreatic insufficiency | gram-negative bacteria | NTM (MAC or non-MAC) | 0.712 |
| a/c pancreatitis | family history | gram-negative bacteria | 0.708 |
| family history | bronchiectasis and/or tree-in-bud | gram-positive bacteria | 0.701 |

**Fig 2.**
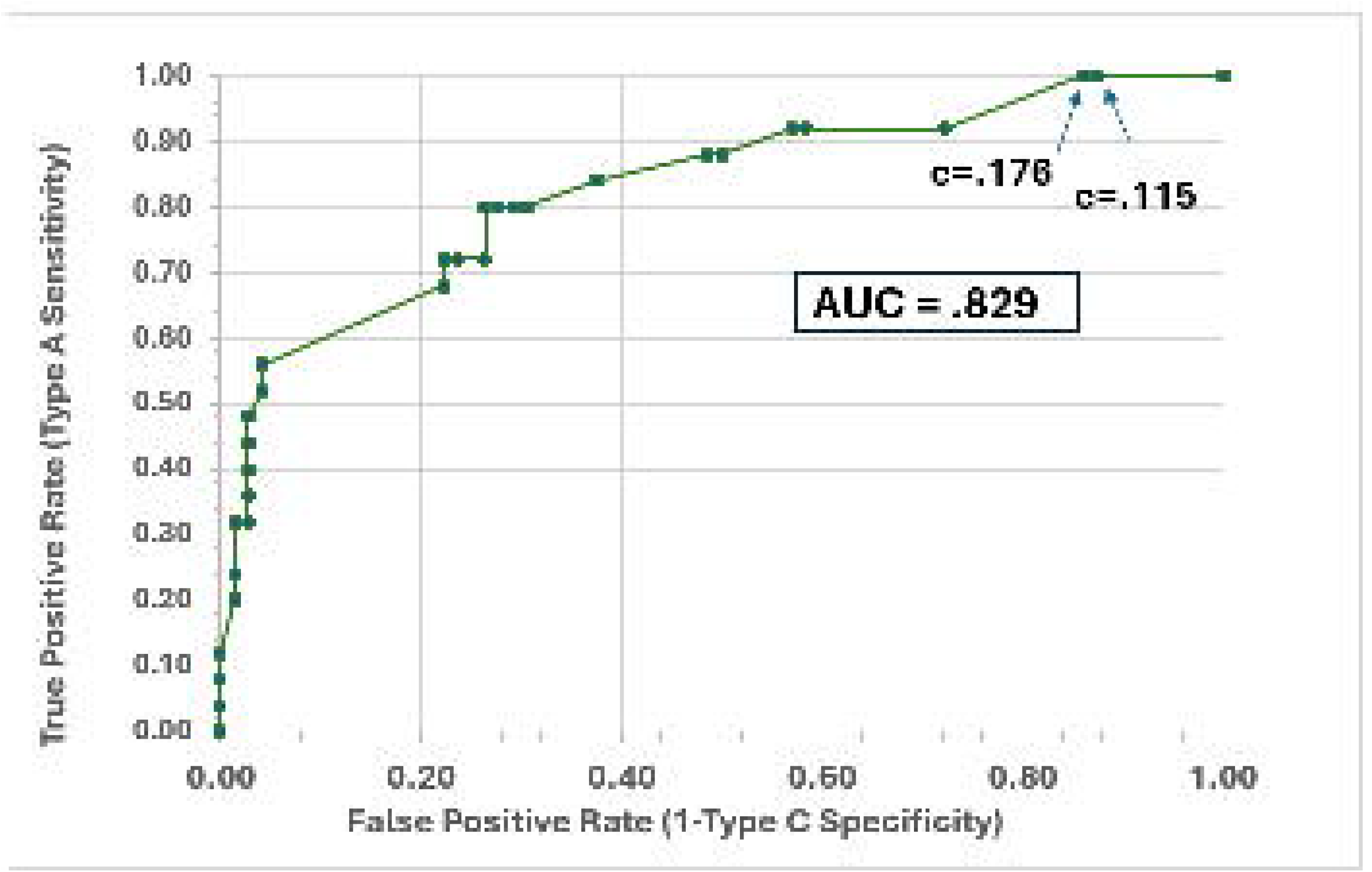
ROC Curve Resulting from Application of Screening Algorithm to Dataset 1

Tables 4 and 5 below show the sensitivity and specificity values obtained by applying the screening model with the decision rule that a patient is recommended for CF testing only if Criterion 1 is satisfied. Table 4 applies to Dataset 1 and Table 5 applies to Dataset 2. The number (n) of Dataset 1 patients is indicated for Type *A, B*, and *C*. The *C*_0_ value of 0.115 is ruled out because the screening algorithm would recommend all 171 Dataset 1 patients for CF testing. As a result, the choice *C*_0_=0.176 is made.

**Table 4.** Dataset 1 Sensitivity and Specificity Results.

| *C*<sub>0</sub> Value | Type A (n=25) | Type *B*<sub>1</sub> (n=24) | Type *B*<sub>0</sub> (n=50) | Type C (n=72) |
| --- | --- | --- | --- | --- |
|  | Sensitivity | Sensitivity | Sensitivity | Specificity |
| 0.176 | 1.000 | 0.958 | 0.840 | 0.139 |
| 0.115 | 1.000 | 1.000 | 1.000 | 0.000 |

**Table 5.** Dataset 2 Sensitivity and Specificity Results.

| *C*<sub>0</sub> Value | Type A | Type *B*<sub>1</sub> | Type *B*<sub>0</sub> | Type C |
| --- | --- | --- | --- | --- |
|  | Sensitivity | Sensitivity | Sensitivity | Specificity |
| 0.176 | 1.000 | 0.846 | 0.857 | 0.214 |

For both Dataset 1 and Dataset 2, the screening model was next applied with the decision rule that a patient is recommended for CF testing if either Criterion 1 or Criterion 2 is satisfied. It is then the case that for both datasets all of the Type *B*_*1*_ and all of the Type *B*_*0*_ patients are recommended for CF testing. So, in both cases, the sensitivity values increase to 1.00. In addition, for both Dataset 1 and Dataset 2, the specificity value for Type *C* decreases significantly (to zero in the case of Dataset 1 and to.036 in the case of Dataset 2). Both databases include only those Type *C* patients recommended for CF testing based on clinical judgment. Without the inclusion in the database of Type *C* patients not recommended for CF testing, the calculated value for the screening model Type *C* specificity is understated. The application of the bootstrap method to the screening model to quantify the error in the estimated Type *C* specificity value requires knowledge of the CF classifications of the patients not recommended for CF testing as well as knowledge of the CF classifications of the patients recommended for CF testing. Since the CF classifications of the patients not recommended for CF testing based on clinical judgment are unknown, the bootstrap method was not implemented.

## 4. Discussion

This pilot study demonstrates the feasibility of developing a screening model based on clinical characteristics to assist clinicians in determining which patients to test for CF. Using a set of selected clinical characteristics, the screening model described can be reliably used to recommend for CF testing those patients that testing will classify as CF (with a sensitivity of 1), as well as patients that CF testing will classify as CFNR (also with a sensitivity of 1).

Late diagnosed CF is heterogeneous with presentations other than those that are “classic” and needs to be considered in all ages, races, and ethnicities. It is important to consider the diagnosis of CF or CFNR/CFTR-RD in all patients with compatible clinical features. However, many of these features are not sensitive or specific to CF.(12) With the advent of CFTR modulator therapy, increasing numbers of adults with bronchiectasis coming to medical attention in the community, limited centers with concentrated expertise in CF/non-CF bronchiectasis, and rising healthcare costs, it is important to develop a framework to assist in the determination of which patients should be referred for CF genetic and sweat chloride testing.

Datasets 1 and 2 both indicate that patients satisfying Criterion 2 are either Type B or Type C. Discriminating between these two possibilities requires that each patient satisfying Criterion 2 be recommended for CF testing. Recommending for CF testing each patient satisfying Criterion 2 is key to the prediction model achieving a Type B sensitivity value of 1.000.

With further testing, the screening model described can be developed into a computer application with a user-friendly interface that is immediately accessible to clinicians with varying levels of experience in the diagnosis of CF. This application would supplement clinical judgment in determining when sweat chloride and CFTR genetic mutation testing are appropriate. The authors believe this to be the first study to demonstrate a systematic approach in using clinical features to create such a screening model. Prior work has suggested the use of combinations of sweat chloride, genetic screening, and nasal potential difference (NPD) in the diagnosis of CF (13) among adults with bronchiectasis. However, sweat chloride and NPD testing are not universally available. Therefore, the use of a practical model incorporating commonly obtainable diagnostic elements to determine which patients to send for these tests will be of significant clinical utility. Furthermore, because a significant number of adult patients ultimately diagnosed with CF or CFTR-RD have atypical variants and borderline sweat chloride levels, a computer application that uses the entire clinical picture will prove valuable in the diagnosis.

As indicated in the diagnostic flow diagram in Figure 1, the first step in diagnosis of CF is to obtain a sweat test. As a minimally invasive and relatively inexpensive test, the threshold to obtain sweat tests in adults with the outlined unexplained symptoms should be very low. However, if sweat test results are intermediate (30-59) or diagnostic of CF (>60), or if CF is still felt to be a possibility despite normal sweat test results, with no clear alternative diagnoses, then it is appropriate to pursue complete CFTR gene sequencing with the inclusion of intronic deletions. Limited CFTR variant panels are going to be of less value in this population with higher probability of rare variants. Making sweat testing more accessible in all geographic settings (and potentially remote testing in the future) may have a significant impact on provider/patient willingness to complete testing. The cost of missing a CF diagnosis is not acceptable, and late diagnoses of CF are difficult to make, even for CF experts. Referral to a CF program should be best practice with diagnostic uncertainty and hopefully can be accomplished more easily with the availability of telehealth.

This study has several strengths. Importantly, it introduces and takes the first steps of early validation of an algorithm that uses clinical variables to determine the likelihood of CF. The algorithm is based on real-world data which is available to most clinicians, irrespective of their resources and comfort with diagnosing CF.

The algorithm is also developed with deliberate efforts to focus on predictive clinical factors. An illustrative example of a patient where our prediction model is applied is in the supplement S.5.

## 5. Limitations

This study has several notable limitations. First, it is a relatively small, single-center study, and the screening model requires wider external validation before it can be put into widespread clinical use. Second, the clinical determination of CF vs CFNR(includes CFTR-RD) vs control, while made by an experienced and qualified CF specialist, was not externally adjudicated. Since CFTR variant analysis was not done for every Type *B*_0_ or *C* patient, and ultimately the diagnosis of CF was based on clinical parameters as assessed by the CF center director (RB), there is risk of CF misclassification. Third, the selected population is subject to referral bias. This is a retrospective study, such that for each dataset patient, clinical judgment was used to decide on whether patients were sent for sweat testing. Thus, excluded from the dataset are all Type *A, B*_1_, and *B*_0_ patients who would not be referred for CF testing based on clinical judgment but would be referred for CF testing by the prediction model. Also, many if not most Type C patients who were not referred for CF testing based on clinical judgment also would not have been referred by the prediction model, thus lowering specificity of the model. Consequently, actual CF classifications are unknown for those patients who are not referred for CF testing based on clinical parameters. Finally, although only patients with adequate clinical information were included, we cannot exclude the possibility that some clinical information (such as imaging findings and culture data) was incomplete because it was not included in the EMRs used for data collection. Clinical variables assumed to be highly likely absent based on clinical chart review were considered absent and may have been misclassified. However, it is the judgment of the authors that the percent of misclassifications is likely to be low. Addressing all of these limitations would require a prospective study design such that each dataset patient is tested for CF whether or not CF testing is supported by clinical judgment.

## 6. Conclusions

Our pilot study achieved the objective of designing, implementing, and testing a predictive model to screen adult patients for CF testing. The study results indicate that for both Dataset 1 and Dataset 2, the screening model matched the performance of clinical judgment in recommending both all Type *A* and all Type *B* patients for CF testing. In addition, the screening model converted to true negatives 1.6% (2/128) of the Type *C* patients who were false positives based on clinical judgment. In summary, an algorithm based on predictive clinical factors for the diagnosis of CF and CFNR is feasible and reproducible and should be further validated in a prospective multicenter study to evaluate its potential utility in determining which patients merit testing for CF.

## Data Availability

Dryad Data Repository https://datadryad.org/ DOI: 10.5061/dryad.tht76hfcz

https://doi.org/10.5061/dryad.tht76hfcz

## Abbreviation List

ABPA: Allergic bronchopulmonary aspergillosis
A/c: Acute, recurrent and/or chronic
AUC: Area under receiver operating characteristic curve
BCC: *Burkholderia cepacia* complex
BMI: Body mass index
CF: Cystic Fibrosis
CFNR: Cystic Fibrosis Not Resolved
CFTR: Cystic fibrosis transmembrane regulator
CFTR-RD: Cystic Fibrosis Transmembrane regulator-related disorder
CI: Confidence interval
CT: Computerized tomography
C_0_: ROC curve cutoff probability used in determining whether to recommend CF testing.
D: Dataset 1, Development dataset
Expanded: Panel of CFTR mutation analysis of at least 200 CF variants
FEV-1: Forced expiratory volume in one second
IQR: Interquartile range
LR: Logistic regression
M: *Mycobacterium*
MAC: *Mycobacterium avium* complex
ML: Maximum likelihood
MRSA: Methicillin resistant *Staphylococcus aureus*
MSSA: Methicillin sensitive *Staphylococcus aureus*
Not: tested No CFTR mutation analysis performed
Non-Mac: All other mycobacterial species other than *mycobacterium avium complex*
NTM: Nontuberculous *mycobacterium*
PPV: Positive predictive value
ROC: Receive operator curve
SP: Sample proportion
Specific: 10 or less CFTR variants tested
Spp.: Species Standard Panel of CFTR mutation analysis between 23-16 variants
TP: True positive
V: Dataset 2, Validation dataset
VVCC: Variant of varying clinical significance

## 7. Author Contributions

RB is the guarantor of the manuscript; he had full access to the data from the study at all times and takes responsibility for the integrity of all data and analyses. RB and BB contributed to study design.

BB performed the data analysis. RB, YM, and BB wrote the first draft of the manuscript. RB and BB wrote the final draft of the manuscript. All authors approved the final version of the manuscript. The preliminary results of this study were presented in abstract form at the North American Cystic Fibrosis Conference in November 2021 and September 2024. There are no financial or non-financial disclosures. There were no sponsors of this study.

## 8. Acknowledgements

Robert Wright, MD, Joe Carmona, BS, Cori Stritzel, RN, Dominque Rosuello, Mony Son, MD, Ginerosa Carbone, MD, Austin Moser, MD, and Amelia Jones, MS, CCRP for preparation of the manuscript.

