## Supplement for "Pilot screening multivariable prediction model for late diagnosed cystic fibrosis"

**Application of Multi-Model Logistic Regression Analysis**

**to Screening for CF Testing**

**S.1 Description of Multi-Model Logistic Regression Analysis.** For each of the 7 CF-related clinical conditions listed in Table 2, the associated indexed binary indicator variable (denoted by *I_k_*) when applied to a patient takes the value 1 if the patient is positive for the clinical condition *k* and takes the value 0 if the patient is negative for clinical condition *k*. Let *P_A_* denote the probability that CF testing will assign a given patient the Type *A* CF classification. The assumption is made that for any specified subset of 1, 2, or 3 of the 7 CF-related clinical conditions for which the patient diagnosis is known, the patient odds ratio *P_A_* / (1- *P_A_*) can be approximated as a log-linear function of the indicator variables that correspond to the specified CF-related clinical conditions.

The motivation for limiting the size of the subset of indicator variables to 3 is to avoid the large errors in the estimated values for the LR model parameter likely to occur if a single 7-variate LR model were used to estimate *S_A_*. Limiting the number of indicator variables in each LR equation to 3 is acceptable because *S_A_* is taken to be the largest of the *P_A_* values for all of the 35 subsets of size 3 of the set of 7 CF-related clinical conditions. *S_A_* is viewed as a numerical measure of the strength of the available diagnostic evidence provided by the 7 CF-related clinical conditions supporting a patient CF classification of Type *A*.

**S.2 Estimation of the Parameter Values for Each of the Multiple LR Models.** As indicated**,** the number of 3-variate LR models is 35 (the number of distinct subsets of size 3 that can be drawn from a set of size 7). Similarly, there are 21 bivariate LR models and 7 univariate LR models. The total number of LR models is therefore 63. Using Dataset 1, the Generalized Reduced Gradient (GRG) nonlinear optimization method is used to determine the maximum likelihood estimates of the parameter values for each of these 63 LR models. The following is a description of the procedure used to calibrate to Dataset 1 each of the LR models used by the prediction model.

- For a given patient, each of 35 3-variate logistic regression models provides an estimate for the probability that the patient is Type A.
- Each variate in an LR model indicates either a positive diagnosis (variate value = 1) or a negative diagnosis (variate value = 0) for the clinical condition associated with that variate.
- Each LR model variate for which the associated clinical condition has a positive diagnosis contributes positively to the LR model estimate for the probability that the patient is Type A.
- Each LR model variate for which the associated clinical condition has a negative diagnosis contributes neither positively nor negatively to the LR model estimate for the probability that the patient is Type A.
- As indicated, maximum likelihood estimation is used to fit the values of the parameters of each LR model to Database 1.
- In calibrating the LR models, if Database 1 records a patient diagnosis for a clinical condition as unknown, the assumption is made that the patient was not tested for that clinical condition because it was clinically judged that the resulting diagnosis would very likely be negative.
- In calibrating the LR models, each diagnosis recorded as unknown is therefore treated as a negative diagnosis. Consequently, the clinical condition data for each of the 171 Database 1 patients is used in calibrating each of the LR models

**S.3 Screening Model Predictive Performance:** To test screening model predictive performance, the predictive model was first applied to Dataset 1, the dataset used to calibrate the LR model parameters. This generated values for the following predictive performance metrics: (1) the Type *A* sensitivity, (2) the Type *B_1_* sensitivity, (3) the Type *B_0_* sensitivity, and (4) the Type *C* specificity, The values obtained for these four metrics are recorded in Table 4

The second step in evaluating the screening model predictive applied the screening to Dataset 2, collected over a time period subsequent to that for the 171-patient study database and based on an entirely different set of patients. The values obtained for the four indicated metrics are recorded in Table 5.

**S.4 Application of the LR Models to Patients.** If a 3-variate LR model is applied to a patient, then for each of the CF-related clinical conditions associated with the model, there are 3 possibilities: (1) the patient has been diagnosed as positive for the condition, (2) the patient has been diagnosed as negative for the condition, or (3) it is unknown whether the patient is positive for the clinical condition or negative for the clinical condition. Since the LR model considers 3 CF-related clinical conditions, there are 3^3^ = 27 cases to consider in determining all of the possible values for *P_A_*.

Consider the application of a specific 3-variate LR model to a patient. If the patient diagnosis (positive or negative) is known for each of the CF-related conditions associated with the 3-variate LR model, then the 3-variate LR model being applied to the patient is used to determine the estimate *P_A_*. If the patient diagnosis is known for only 2 of the CF-related clinical conditions associated with the 3-variate LR model, then the bivariate model based on those 2 CF-related clinical conditions is used to determine the estimate *P_A_*. If the patient diagnosis is known for only 1 of the CF-related clinical conditions associated with the 3-variate LR model, then the univariate model based on that single CF-related clinical condition is used to determine the estimate *P_A_*. Finally, if the patient diagnosis is unknown for each of the 3 CF-related clinical conditions associated with the 3-variate LR model, then *P_A_* is taken to be the observed fraction of the Dataset 1 patients that are classified as Type *A*. The required values to apply the LR analysis are stored in 35 precomputed tables.

In applying the LR analysis to a particular patient, a lookup is executed for each table to find the appropriate input values for the given patient. The score *S_A_* assigned to the patient is then taken to be the largest of the 35 calculated *P_A_* values. The decision rule, as described in the Methods section, is then applied to determine whether the screening model recommendation should be to test the patient for CF.

**S.5 Illustrative example**:

The patient is between 51 to 60 yrs old (male) with mild bronchiectasis and tree-in-bud opacities, recurrent growth of MSSA, no other bacterial infections, no NTM, pancreatic sufficiency, and no history of pancreatitis.

The numerical score *S_A_* calculated by the prediction model for this patient is 0.407. Since *S_A_* ≥.176 the patient score is sufficient that the prediction model would recommend CF testing for this patient. The actual CF testing results revealed a sweat test of 71/68, and the 97 CFTR mutation analysis and expanded CFTR variant analysis through the Cystic Fibrosis Foundation Mutation Analysis Program (CFFMAP) revealed single pathogenic variant F508del. The patient was diagnosed with CF.

**Supplementary Data Tables**

**DEVELOPMENT COHORT – DATASET 1**

**TABLE S1-D: DEMOGRAPHICS**

| **CF Classification** | **Type *A* (CF)** | | | **Type *B* (CFNR)** | | | | **Type *C* (Control)** | | | | **Total** | | |
| --- | --- | --- | --- | --- | --- | --- | --- | --- | --- | --- | --- | --- | --- | --- |
| ***Subjects* (n, %)** | 25 | | 15% | 74 | | 43% | | 72 | | 42% | | 171 | |  |
| ***Age* (n)** |  |  |  |  |  |  |  |  |  |  |  |  |  |  |
| **Median (yrs)** | 53 | |  | 61 | |  | | 58 | |  | | 59 | |  |
| **IQR (yrs)** | 31 | |  | 24 | |  | | 32 | |  | | 30 | |  |
| ***Sex* (n, %)** |  |  |  |  |  |  |  |  |  |  |  |  |  |  |
| **Male** | 8 | | 32% | 20 | | 27% | | 22 | | 31% | | 50 | | 29% |
| **Female** | 17 | | 68% | 54 | | 73% | | 50 | | 69% | | 121 | | 71% |
| ***Sweat Cl* (n, %)** |  |  |  |  |  |  |  |  |  |  |  |  |  |  |
| **≥60 mmol/L** | 20 | | 80% | 4 | | 5% | | 0 | | 0% | | 24 | | 14% |
| **30-59 mmol/L** | 4 | | 16% | 58 | | 78% | | 1 | | 1% | | 63 | | 37% |
| **≤29 mmol/L** | 1 | | 4% | 12 | | 16% | | 71 | | 99% | | 84 | | 49% |
| ***CFTR Variant Analysis* (n, %)** |  |  |  |  |  |  |  |  |  |  |  |  |  |  |
| **Patients Tested** | 25 | | 100% | 63 | | 85% | | 34 | | 47% | | 122 | | 71% |
| **Mutations=0** | 7 | | 28% | 34 | | 54% | | 34 | | 100% | | 75 | | 61% |
| **Mutations=1** | 4 | | 16% | 28 | | 44% | | 0 | | 0% | | 32 | | 26% |
| **Mutations=2** | 13 | | 52% | 1 | | 2% | | 0 | | 0% | | 14 | | 11% |
| **Mutations=3** | 1 | | 4% | 0 | | 0% | | 0 | | 0% | | 1 | | 1% |
| ***FEV 1* (L, % predicted)** |  |  |  |  |  |  |  |  |  |  |  |  |  |  |
| **Median** | 2.59 | | 90% | 2.05 | | 77% | | 2.10 | | 81% | | 2.07 | | 81% |

**TABLE S2-D: CFTR TESTING PERFORMED**

| **CF Classification** | **Type *A*** | | **Type *B*** | | **Type *C*** | | **Total** | | |
| --- | --- | --- | --- | --- | --- | --- | --- | --- | --- |
| **Subjects (n*, %)*** |  |  |  |  |  |  |  |  |  |
| **Standard (23-106)** | 13 | 52% | 29 | 39% | 34 | 47% | | 76 | 44% |
| **Expanded (200+)** | 10 | 40% | 32 | 43% | 0 | 0% | | 43 | 25% |
| **Specific (< 10)** | 2 | 8% | 1 | 1% | 1 | 1% | | 3 | 2% |
| **Not Tested** | 0 | 0% | 11 | 15% | 37 | 51% | | 48 | 28% |
| **Unknown Type of Testing** | 0 | 0% | 1 | 1% | 0 | 0% | | 1 | 1% |

**TABLE S3-D: BREAKDOWN OF TYPE *B* PATIENTS (CFNR)**

| **Type *B*_0_ and**  **Negative CFTR Testing** | **Type *B*_0_ and**  **No CFTR Testing Performed** | **Type *B*_0_ with non-CF causing variants** | **Type *B*_1_** |
| --- | --- | --- | --- |
| 34 | 11 | 2 | 26 |

**TABLE S4-D: CF-RELATED CLINICAL CHARACTERISTICS**

| **CF Classification** | **Type *A*** | | **Type *B*** | | **Type *C*** | | **Total** | |
| --- | --- | --- | --- | --- | --- | --- | --- | --- |
| **Subjects (n, %)** | 25 | | 74 | | 72 | | 171 | |
| **Pancreatic Insufficiency** | 8 | 32% | 3 | 4% | 1 | 1% | 12 | 7% |
| **Acute/Chronic Pancreatitis** | 4 | 16% | 5 | 7% | 2 | 3% | 11 | 6% |
| **Infertility** | 4 | 16% | 2 | 3% | 0 | 0% | 6 | 4% |
| **Family History** | 7 | 28% | 11 | 15% | 1 | 1% | 19 | 11% |
| **Bronchiectasis** | 12 | 48% | 50 | 68% | 45 | 63% | 107 | 63% |
| **Tree-in Bud Opacities** | 8 | 28% | 29 | 39% | 30 | 42% | 67 | 39% |
| **Bronchiectasis and/or Tree-in-Bud Opacities** | 14 | 56% | 53 | 72% | 48 | 67% | 115 | 67% |
| **Acute/Chronic Sinusitis** | 9 | 36% | 30 | 41% | 36 | 50% | 75 | 44% |
| **Nasal Polyps** | 1 | 4% | 12 | 16% | 13 | 18% | 26 | 15% |
| **Pseudomonas aeruginosa** | 8 | 32% | 25 | 34% | 27 | 38% | 60 | 35% |
| **MRSA** | 6 | 24% | 6 | 8% | 8 | 11% | 20 | 12% |
| **MSSA** | 7 | 28% | 14 | 19% | 19 | 26% | 40 | 23% |
| **Aspergillus species** | 5 | 20% | 10 | 14% | 13 | 18% | 28 | 16% |
| **Stenotrophomonas maltophilia** | 1 | 4% | 5 | 7% | 6 | 8% | 12 | 7% |
| **NTM (MAC)** | 4 | 16% | 24 | 32% | 13 | 17% | 41 | 24% |
| **NTM (non-MAC)** | 2 | 8% | 8 | 11% | 8 | 11% | 18 | 11% |
| **NTM (MAC and/or non-MAC)** | 5 | 20% | 28 | 38% | 16 | 22% | 49 | 29% |
| **M. abscessus complex** | 2 | 8% | 2 | 1% | 0 | 0% | 4 | 2% |
| **M. chelonae** | 0 | 0% | 0 | 0% | 1 | 1% | 1 | 1% |
| **M. fortuitum** | 0 | 0% | 0 | 0% | 2 | 3% | 2 | 1% |
| **M. gordonae** | 0 | 0% | 2 | 3% | 1 | 1% | 3 | 2% |
| **M. lentiflavum** | 0 | 0% | 1 | 1% | 0 | 0% | 1 | 1% |
| **M. mucogenicum** | 0 | 0% | 0 | 0% | 1 | 1% | 1 | 1% |
| **M. xenopi** | 0 | 0% | 1 | 1% | 1 | 1% | 2 | 1% |
| **M. kumamotonense** | 0 | 0% | 0 | 0% | 1 | 1% | 1 | 1% |
| **Alcaligenes xylosoxidans** | 0 | 0% | 3 | 4% | 2 | 3% | 5 | 3% |
| **Burkholderia cepacia complex** | 1 | 4% | 0 | 0% | 0 | 0% | 1 | 1% |
| **Wangiella dermatitidis** | 0 | 0% | 0 | 0% | 0 | 0% | 0 | 0% |
| **Haemophilus influenza** | 5 | 20% | 14 | 19% | 15 | 21% | 34 | 20% |
| **Streptococcus pneumoniae** | 2 | 8% | 1 | 1% | 5 | 7% | 8 | 5% |
| **Moraxella catarrhalis** | 0 | 0% | 0 | 0% | 4 | 6% | 4 | 2% |
| **Asthma** | 11 | 44% | 29 | 39% | 30 | 42% | 70 | 41% |
| **ABPA** | 2 | 8% | 7 | 9% | 4 | 6% | 13 | 8% |
| **Diabetes Mellitus** | 3 | 12% | 3 | 4% | 3 | 4% | 9 | 5% |
| **Gram Negative Infections** | 13 | 52% | 35 | 47% | 39 | 54% | 87 | 51% |
| **Gram Positive Infections** | 12 | 48% | 20 | 27% | 21 | 29% | 53 | 31% |

**TABLE S5-D: CFTR Variants TESTED**

| **CF Classification** | **Type *A*** | **Type *B*** |
| --- | --- | --- |
| **Variants(s)** | **# Patients** | **# Patients** |
| **1716G>A** |  | 1 |
| **178G>T** |  | 2 |
| **2789+5G>A / A455V (unknown)** | 2 |  |
| **2789+5G>A** |  | 2 |
| **5T;TG11 (non-CF causing)** |  | 2 |
| **5T;TG12 (VVCC)** |  | 3 |
| **F508∆ / A455E** | 1 |  |
| **A455V(unknown)/ 2789+5G> / Q372L (unknown)** | 1 |  |
| **1046C>T (VVCC** |  | 1 |
| **2183AA>G** |  | 1 |
| **274-6T>C (VVCC)** | 1 |  |
| **D1152H(VVCC) / R1162X** | 1 |  |
| **F311L** | 1 |  |
| **F508∆** |  | 11 |
| **F508∆ / 2789+5G>A** | 1 |  |
| **F508∆ / 5T;TG12 (VVCC)** | 1 |  |
| **F508∆ / c.1457C>T** | 1 |  |
| **F508∆ / L206W** | 1 |  |
| **F508∆ / R117H (VVCC)** | 4 |  |
| **F575Y / 1874insT** | 1 |  |
| **L997F (VVCC)** |  | 1 |
| **G542X** | 1 |  |
| **N1303K** |  | 1 |
| **R117H (VVCC)/ c.1584G>A (non-CF causing)** |  | 1 |
| **R117H (VVCC)** |  | 2 |
| **R347P** | 1 |  |

**VALIDATION COHORT – DATASET 2**

**TABLE S1-V: DEMOGRAPHICS**

| **CF Classification** | **Type *A* (CF)** | | **Type *B* (CFNR)** | | **Type *C* (Control)** | | **Total** | |
| --- | --- | --- | --- | --- | --- | --- | --- | --- |
| ***Subjects* (n, %)** | 4 | 5% | 27 | 31% | 56 | 64% | 87 |  |
| ***Age* (n)** |  |  |  |  |  |  |  |  |
| **Median (yrs)** | 61.0 |  | 61.0 |  | 62.5 |  | 62.0 |  |
| **IQR (yrs)** | 25.3 |  | 38.5 |  | 27.0 |  | 30.0 |  |
| ***Sex* (n, %)** |  |  |  |  |  |  |  |  |
| **Male** | 1 | 25% | 11 | 41% | 16 | 29% | 28 | 32% |
| **Female** | 3 | 75% | 16 | 59% | 40 | 71% | 59 | 68% |
| ***Sweat Cl* (n, %)** |  |  |  |  |  |  |  |  |
| **≥60 mmol/L** | 3 | 75% | 0 | 0% | 0 | 0% | 3 | 3% |
| **30-59 mmol/L** | 1 | 25% | 16 | 59% | 3 | 5% | 20 | 23% |
| **≤29 mmol/L** | 0 | 0% | 11 | 41% | 53 | 97% | 64 | 74% |
| ***CFTR Mutation Analysis* (n, %)** |  |  |  |  |  |  |  |  |
| **Patients Tested** | 4 | 100% | 24 | 89% | 18 | 32% | 46 | 53% |
| **Mutations=0** | 2 | 50% | 11 | 46% | 17 | 94% | 30 | 65% |
| **Mutations=1** | 0 | 0% | 13 | 54% | 1 | 6% | 14 | 30% |
| **Mutations=2** | 2 | 50% | 0 | 0% | 0 | 0% | 2 | 4% |
| ***FEV 1* (L, % predicted)** |  |  |  |  |  |  |  |  |
| **Median** | 2.14 | 72% | 2.51 | 93% | 2.43 | 84% | 2.44 | 89% |

**TABLE S2-V: CFTR TESTING PERFORMED**

| **CF Classification** | **Type *A*** | | **Type *B*** | | **Type *C*** | | **Total** | |
| --- | --- | --- | --- | --- | --- | --- | --- | --- |
| **Subjects (n*, %*)** |  |  |  |  |  |  |  |  |
| **Standard (23-106)** | 3 | 75% | 16 | 59% | 18 | 32% | 37 | 43% |
| **Expanded (200+)** | 0 | 0% | 6 | 22% | 0 | 0% | 6 | 7% |
| **Specific (< 10)** | 0 | 0% | 0 | 0% | 0 | 0% | 0 | 0% |
| **Not Tested** | 0 | 0% | 3 | 11% | 38 | 68% | 41 | 47% |
| **Unknown Type of Testing** | 1 | 25% | 2 | 7% | 0 | 0% | 3 | 3% |

**TABLE S3-V: BREAKDOWN OF TYPE *B* PATIENTS (CFNR)**

| **Type *B*_0_ and**  **Negative CFTR Testing** | **Type *B*_0_ and**  **No CFTR Testing Performed** | **Type *B*_0_ with non-CF causing mutations** | **Type *B*_1_** |
| --- | --- | --- | --- |
| 11 | 2 | 2 | 12 |

**TABLE S4-V: CF-RELATED CLINICAL CHARACTERISTICS**

| **CF Classification** | **Type *A*** | | **Type *B*** | | **Type *C*** | | **Total** | |
| --- | --- | --- | --- | --- | --- | --- | --- | --- |
| **Subjects (n, %)** | 4 | | 27 | | 56 | | 87 | |
| **Pancreatic Insufficiency** | 2 | 50% | 3 | 11% | 1 | 2% | 6 | 7% |
| **Acute/Chronic Pancreatitis** | 0 | 0% | 6 | 22% | 2 | 4% | 8 | 9% |
| **Infertility** | 0 | 0% | 0 | 0% | 0 | 0% | 0 | 0% |
| **Family History** | 0 | 0% | 3 | 11% | 2 | 4% | 5 | 6% |
| **Bronchiectasis** | 4 | 100% | 17 | 63% | 32 | 57% | 53 | 61% |
| **Tree-in Bud Opacities** | 1 | 25% | 8 | 30% | 17 | 30% | 26 | 30% |
| **Bronchiectasis and/or Tree-in-Bud Opacities** | 4 | 100% | 17 | 63% | 33 | 59% | 54 | 62% |
| **Acute and/or Chronic Sinusitis** | 3 | 75% | 9 | 33% | 39 | 70% | 51 | 59% |
| **Nasal Polyps** | 1 | 25% | 5 | 19% | 16 | 29% | 22 | 25% |
| **Pseudomonas aeruginosa** | 1 | 25% | 9 | 33% | 19 | 34% | 29 | 33% |
| **MRSA** | 0 | 0% | 1 | 4% | 1 | 2% | 2 | 2% |
| **MSSA** | 2 | 50% | 10 | 37% | 6 | 11% | 18 | 21% |
| **Aspergillus species** | 1 | 25% | 4 | 15% | 13 | 23% | 18 | 21% |
| **Stenotrophomonas maltophilia** | 1 | 25% | 5 | 19% | 4 | 7% | 10 | 11% |
| **NTM (MAC)** | 1 | 25% | 2 | 7% | 12 | 21% | 15 | 17% |
| **NTM (non-MAC)** | 2 | 50% | 3 | 11% | 3 | 5% | 8 | 9% |
| **NTM (MAC and/or non-MAC)** | 3 | 75% | 4 | 15% | 14 | 25% | 21 | 24% |
| **M. abscessus complex** | 0 | 0% | 0 | 0% | 2 | 4% | 2 | 2% |
| **M. chelonae** | 0 | 0% | 0 | 0% | 2 | 4% | 2 | 2% |
| **M. gordonae** | 2 | 50% | 2 | 7% | 0 | 0% | 4 | 5% |
| **M. lentiflavum** | 0 | 0% | 0 | 0% | 1 | 1% | 1 | 1% |
| **M. kansasii** | 0 | 0% | 1 | 4% | 0 | 0% | 1 | 1% |
| **Alcaligenes xylosoxidans** | 0 | 0% | 0 | 0% | 0 | 0% | 0 | 0% |
| **Burkholderia cepacia complex** | 0 | 0% | 0 | 0% | 0 | 0% | 0 | 0% |
| **Wangiella dermatitidis** | 0 | 0% | 0 | 0% | 0 | 0% | 0 | 0% |
| **Haemophilus Influenza** | 1 | 25% | 1 | 4% | 9 | 16% | 11 | 13% |
| **Streptococcus pneumoniae** | 0 | 0% | 2 | 7% | 5 | 9% | 7 | 8% |
| **Moraxella catarrhalis** | 0 | 0% | 0 | 0% | 1 | 2% | 1 | 1% |
| **Asthma** | 0 | 0% | 17 | 63% | 41 | 73% | 58 | 67% |
| **ABPA** | 2 | 50% | 3 | 11% | 5 | 9% | 10 | 11% |
| **Diabetes Mellitus** | 1 | 25% | 3 | 11% | 3 | 5% | 7 | 8% |
| **Gram Negative** | 2 | 50% | 10 | 37% | 28 | 50% | 40 | 46% |
| **Gram Positive** | 2 | 50% | 10 | 37% | 6 | 11% | 18 | 21% |

**TABLE S5-V: CFTR MUTATIONS TESTED**

| **CF Classification** | **Type *A*** | **Type *B*** | **Type C** |
| --- | --- | --- | --- |
| **Mutation(s)** | **# Patients** | **# Patients** | **# Patients** |
| **2988G>A** |  | 1 |  |
| **5T;TG11(non CF causing)** |  | 2 | 1 |
| **1766+1G>A** |  | 1 |  |
| **D1152H (VVCC)** |  | 1 |  |
| **F508∆** |  | 7 |  |
| **F508∆ / G542X** | 1 |  |  |
| **G542X /D1152H (VVCC)** | 1 |  |  |
| **R117H (VVCC)** |  | 1 |  |
